# Hypertension Pharmacotherapy in Skilled Nursing Facilities: A Real-World Evidence Study

**DOI:** 10.64898/2026.07.21.26358585

**Authors:** Huda Ashraf, Katherine E. Mathers, Brittin Wanger, Tyler M. Saumur

## Abstract

**Objectives:** To evaluate rates of pharmacological hypertension orders and identify resident- and facility-level predictors of pharmacologic care among skilled nursing facility (SNF) residents in the United States.

**Design:** Retrospective, observational study.

**Setting and Participants:** Electronic Health Record data from 1,285,062 long-term care residents in PointClickCare’s Life Sciences database in facility on April 30, 2025 were reviewed, and 553,519 SNF residents with a documented hypertension diagnosis were identified.

**Methods:** The presence and absence of medication orders for antihypertensive treatment recommended by the International Society of Hypertension was assessed. Descriptive analyses summarized demographic and clinical characteristics, and a modified Poisson regression model was used to estimate risk ratios (RRs) for having a medication order, adjusting for demographic, clinical, and facility characteristics.

**Results:** Overall, 87.7% of residents diagnosed with hypertension had at least one antihypertensive medication order. Calcium channel blockers (44.3%) and beta blockers (43.5%) were the most frequently used classes. RRs ranged from 0.91 to 1.09. Higher likelihoods of antihypertensive orders were observed among residents prescribed hyperlipidemia and diabetes medication (RR = 1.09 and 1.05, respectively), while lower likelihoods of treatment were observed for other payer types (RR = 0.91), diabetes diagnoses (RR = 0.95), and hyperlipidemia diagnoses (RR = 0.98).

**Conclusions and Implications:** Most residents with hypertension had orders for recommended pharmacologic therapy, although important gaps and disparities remain. The predominance of certain medication classes and persistent differences by comorbidity and facility type underscore the need for targeted strategies to improve equitable prescribing and access to evidence-based hypertension management in SNF settings.

## Introduction

Hypertension is among the most common chronic conditions in older adults and is highly prevalent in institutional care. National trends show that among U.S. adults with hypertension, the age-adjusted proportion with controlled blood pressure (<140/90 mmHg) increased from 31.8% in 1999–2000 to 53.8% in 2013–2014 but subsequently declined to 43.7% in 2017–2018.^1^ In long-term care (LTC) settings, overtreatment with hypertension medication is common, with many LTC residents taking multiple antihypertensives and having low blood pressure. ^2–4^ In these frail LTC populations, clinicians must balance potential benefits of blood pressure control against risks associated with intensive antihypertensive therapy, supporting individualized treatment decisions.^5–7^ Indeed, studies have shown that deprescribing antihypertensives can be done relatively safely and effectively when planned and monitored appropriately.^8^

Treatment rates and prescribing patterns for hypertension in LTC vary widely. In a Canadian LTC audit of 733 residents with hypertension (mean age 84 years), 77% were treated and 64% met target blood pressure; most prescribed classes were angiotensin-converting enzyme (ACE) inhibitors (60%), calcium channel blockers (30%), thiazides (24%), and beta-blockers (20%). ^9^ In U.S. nursing homes, 84% of hypertensive residents received at least one antihypertensive agent, with 27.8% treated with beta-blockers despite guideline caution for first-line use in uncomplicated cases. ^10^ A quality improvement review of 75 LTC residents with hypertension aged ≥80 years found 39% of those without comorbidities had hypotensive averages (<130/60 mmHg), raising concerns around overtreatment. ^11^ International Society of Hypertension (ISH) guidelines emphasize individualized therapy, accurate measurement, and risk stratification, recommending essential and optimal standards for older adults, including those in LTC.^12^

Historically documented disparities in hypertension management raise critical questions: what are contemporary treatment rates, and how do they vary by resident characteristics and prescribing patterns? Treatment inequities in antihypertensive and cardiovascular disease management, particularly among older adults in LTC settings, have been linked to older age, frailty, polypharmacy, facility characteristics, and demographic factors.^7^ Untreated or poorly managed hypertension may increase cardiovascular risk, hospitalization, and mortality in frail older adults. Accordingly, this study aimed to evaluate up-to-date patterns of antihypertensive treatment, all medication classes included in current pharmacological hypertension treatment, and several facility–level factors in a large electronic health record (EHR) dataset representative of U.S. older adults in LTC.

## Methods

### Setting and Participants

This retrospective observational study utilized deidentified EHR data from the PointClickCare Life Sciences database, one of the largest real-world datasets of its kind, encompassing clinical information from more than 18 million residents across U.S. LTC and skilled nursing facilities (SNFs). The database includes resident demographics, Minimum Data Set clinical and functional assessments, medication and vaccination records, vital signs, and related data elements. Data is deidentified and expert-determined from residents with facility-obtained consent and covered by business associate agreements with LTC facilities. Exemption from ethics committee oversight was granted by the Research Ethics Boards of the University of Toronto and University of Alberta.

Data were assessed cross-sectionally using a database snapshot of April 30, 2025, with eligible residents required to be active in a skilled nursing facility on that date. EHR data were available for 1,285,062 individuals in LTC facilities. Among these, 553,519 SNF residents had a documented diagnosis of hypertension, identified using ICD-10 code I10. Residents were eligible for inclusion if they had an active hypertension diagnosis. The dataset provided comprehensive information on demographics, comorbidities, prescribed medications, and facility-level characteristics.

### Exposures and Variables of Interest

Residents were categorized based on the presence or absence of medication orders for antihypertensives consistent with the International Society of Hypertension medication classes.^12^ Medication classes included beta blockers, calcium channel blockers, loop diuretics, angiotensin II receptor blockers (ARBs), ACE inhibitors, and alpha-beta blockers.

Variables included demographic characteristics (age, sex, race/ethnicity), insurance status, facility region, and length of stay. Days in facility (DIF) was calculated as the number of days between a resident’s admission and the study end date. Consecutive visits were combined when separated by gaps of seven days or fewer, treating them as a single continuous stay for DIF calculation. Clinical comorbidities such as diabetes, hyperlipidemia, and dementia were included due to their potential influence on prescribing patterns.

### Data Analysis

Data were extracted using SQL and analyzed in Python. Descriptive statistics summarized resident demographics and treatment characteristics. A modified Poisson regression model was used with a log link to estimate prevalence risk ratios (RRs) and 95% confidence intervals (CIs) for having a Type 2 diabetes medication order. Covariates include resident demographics, clinical characteristics, prescription and over-the–counter medication counts, diagnosis counts, and facility characteristics. Categorical variables were converted to binary indicator variables using pre-specified reference categories: female sex, White race, Medicare fee-for-service payer, Southern U.S. region, and non-profit facility ownership. Continuous variables were rescaled prior to model entry to improve interpretability of effect estimates: age was expressed per 10-year increment, BMI per 5-unit increment, number of facility beds per 25-bed increment, and DIF per 30-day increment. Statistical significance was assessed at α = 0.001.

## Results

There were 553,519 residents with hypertension who met the eligibility criteria (Table 1). 485,642 (87.7%) of these residents had orders for antihypertensive medication. Across all hypertensive residents, the mean (SD) age was 76.9 (12.3) years, with the majority of residents being female (59.3%) and White (66%). Residents were distributed across U.S. regions, with the South (37.5%) and Midwest (28.2%) most represented. Primary payer types (at admission) included Medicaid (32.7%), Medicare FFS (28.3%), and Managed Care (24.1%), with smaller proportions covered by private insurance and other sources.

**Table 1.** Demographic and Clinical Characteristics Across Exposure Groups.

|  | **Active Order**<br>**N = 485,642** | **Not Treated**<br>**N = 67,877** | **All Residents**<br>**N = 553,519** | **Order Rate**<br>**87.7%** |
| --- | --- | --- | --- | --- |
| **Age, years** |  |  |  |  |
| Mean (SD) | 77.0 (12.3) | 76.2 (12.9) | 76.9 (12.3) | NA |
| Median (Q1, Q3) | 78 (69, 86) | 77 (68, 86) | 78 (69, 86) | NA |
| Min, Max | 20, 116 | 20, 117 | 20, 117 | NA |
| **Age Category, n (%)** |  |  |  |  |
| ≥90 | 76,971 (15.8%) | 10,242 (15.1%) | 87,213 (15.8%) | 88.3% |
| 85-89 | 67,412 (13.9%) | 8,636 (12.7%) | 76,048 (13.7%) | 88.6% |
| 80-84 | 74,397 (15.3%) | 10,127 (14.9%) | 84,524 (15.3%) | 88.0% |
| 75-79 | 74,875 (15.4%) | 10,107 (14.9%) | 84,982 (15.4%) | 88.1% |
| 70-74 | 65,615 (13.5%) | 9,116 (13.4%) | 74,731 (13.5%) | 87.8% |
| 65-69 | 52,730 (10.9%) | 7,771 (11.4%) | 60,501 (10.9%) | 87.2% |
| 19-64 | 73,642 (15.2%) | 11,878 (17.5%) | 85,520 (15.5%) | 86.1% |
| **Sex, n (%)** |  |  |  |  |
| Female | 290,073 (59.7%) | 38,176 (56.2%) | 328,249 (59.3%) | 88.4% |
| Male | 195,569 (40.3%) | 29,701 (43.8%) | 225,270 (40.7%) | 86.8% |

|  | **Active Order**<br>**N = 485,642** | **Not Treated**<br>**N = 67,877** | **All Residents**<br>**N = 553,519** | **Order Rate**<br>**87.7%** |
| --- | --- | --- | --- | --- |
| **Race, n (%)** |  |  |  |  |
| White | 319,812 (65.9%) | 45,528 (67.1%) | 365,340 (66.0%) | 87.5% |
| Black or African American | 85,397 (17.6%) | 10,086 (14.9%) | 95,483 (17.3%) | 89.4% |
| Unspecified | 55,041 (11.3%) | 8,760 (12.9%) | 63,801 (11.5%) | 86.3% |
| Asian | 10,141 (2.1%) | 1,391 (2.0%) | 11,532 (2.1%) | 87.9% |
| Hispanic or Latino | 9,914 (2.0%) | 1,295 (1.9%) | 11,209 (2.0%) | 88.4% |
| American Indian or Alaska Native | 4,461 (0.9%) | 688 (1.0%) | 5,149 (0.9%) | 86.6% |
| Native Hawaiian or Other Pacific Islander | 876 (0.2%) | 129 (0.2%) | 1,005 (0.2%) | 87.2% |
| **Region, n (%)** |  |  |  |  |
| South | 183,996 (37.9%) | 23,844 (35.1%) | 207,840 (37.5%) | 88.5% |
| Midwest | 137,010 (28.2%) | 19,163 (28.2%) | 156,173 (28.2%) | 87.7% |
| Northeast | 98,053 (20.2%) | 14,770 (21.8%) | 112,823 (20.4%) | 86.9% |
| West | 66,583 (13.7%) | 10,100 (14.9%) | 76,683 (13.9%) | 86.8% |
| **Primary Payer, n (%)** |  |  |  |  |
| Medicaid | 158,397 (32.6%) | 22,533 (33.2%) | 180,930 (32.7%) | 87.5% |
| Medicare FFS | 138,818 (28.6%) | 18,033 (26.6%) | 156,851 (28.3%) | 88.5% |
| Managed Care | 118,695 (24.4%) | 14,886 (21.9%) | 133,581 (24.1%) | 88.9% |
| Private | 54,163 (11.2%) | 8,356 (12.3%) | 62,519 (11.3%) | 86.6% |
| Other | 15,569 (3.2%) | 4,069 (6.0%) | 19,638 (3.5%) | 79.3% |
| **Days in Facility** |  |  |  |  |
| Mean (SD) | 709.7 (790.0) | 691.0 (872.6) | 707.4 (800.6) | NA |
| Median (Q1, Q3) | 429 (98, 1,047) | 331 (67, 968) | 416 (94, 1,039) | NA |
| Min, Max | 2, 3,804 | 2, 3,804 | 2, 3,804 | NA |
| **Stay Type, n (%)** |  |  |  |  |
| Long Stay | 363,895 (74.9%) | 47,396 (69.8%) | 411,291 (74.3%) | 88.5% |
| Short Stay | 121,747 (25.1%) | 20,481 (30.2%) | 142,228 (25.7%) | 85.6% |

|  | **Active Order**<br>**N = 485,642** | **Not Treated**<br>**N = 67,877** | **All Residents**<br>**N = 553,519** | **Order Rate**<br>**87.7%** |
| --- | --- | --- | --- | --- |
| **CCI Score** |  |  |  |  |
| Mean (SD) | 2.8 (2.2) | 2.5 (2.1) | 2.7 (2.2) | NA |
| Median (Q1, Q3) | 2 (1, 4) | 2 (1, 3) | 2 (1, 4) | NA |
| Min, Max | 0, 23 | 0, 20 | 0, 23 | NA |
| **Prescription Medication**<br>**Count** |  |  |  |  |
| Mean (SD) | 9.8 (4.6) | 6.3 (4.3) | 9.4 (4.7) | NA |
| Median (Q1, Q3) | 9 (7, 13) | 6 (3, 9) | 9 (6, 12) | NA |
| Min, Max | 0, 73 | 0, 31 | 0, 73 | NA |
| **Over-The-Counter**<br>**Medication Count** |  |  |  |  |
| Mean (SD) | 7.1 (3.7) | 5.9 (3.8) | 6.9 (3.7) | NA |
| Median (Q1, Q3) | 7 (4, 9) | 6 (3, 8) | 7 (4, 9) | NA |
| Min, Max | 0, 44 | 0, 27 | 0, 44 | NA |
| **Active Diagnosis Count<sup>b</sup>** |  |  |  |  |
| Mean (SD) | 22.5 (10.9) | 21.2 (10.7) | 22.4 (10.9) | NA |
| Median (Q1, Q3) | 20 (15, 28) | 19 (14, 26) | 20 (15, 27) | NA |
| Min, Max | 0, 247 | 0, 189 | 0, 247 | NA |

Abbreviations: CCI=Charlson Comorbidity Index; FFS=fee-for-service; ICD=International Classification of Diseases; Q=quartile; SD=standard deviation.
<sup>a</sup> The maximum age range was capped at 90+ to protect participant privacy, as individuals above 90 years represent a very small subgroup and may be more readily identifiable.
<sup>b</sup> Active diagnoses were based on ICD-10 codes.

Clinical complexity was notable, with a mean Charlson Comorbidity Index (CCI) of 2.7 and residents having an average of 22.4 distinct documented diagnoses. Residents with active hypertension orders were prescribed more medications on average (9.8 vs 6.3 for untreated), while over-the-counter medication use was 7.1 vs 5.9 for untreated. The majority of residents were long-stay, with a median facility stay of 416 days. Calcium channel blocker accounted for the largest proportion of antihypertensive prescriptions among treated residents, utilized in and 44.3% of cases. Beta blockers (43.5%), loop diuretics (36.9%), ACE inhibitors (27%), and angiotensin receptor blockers (25.6%) were also commonly prescribed. Other medication classes were prescribed less frequently, including alpha-beta blockers (1 4.8%), vasodilators (12.5%), thiazide diuretics (9.1%), potassium-sparing diuretics (8.6%), central-acting agents (8.3 %), and alpha blocker (1.7%), with aldosterone antagonists and renin inhibitors prescribed to less than 1% of residents with hypertension (Table 2).

**Table 2.** Active Order Rates by Medication Class.

|  | **Active Order**<br>**N = 485,642** |
| --- | --- |
| **Medication Class, n (%)** |  |
| Calcium Channel Blocker | 215,006 (44.3%) |
| Beta Blocker | 211,458 (43.5%) |
| Loop Diuretics | 179,011 (36.9%) |
| Angiotensin-converting-enzyme Inhibitors | 131,208 (27%) |
| Angiotensin Receptor Blockers | 124,089 (25.6%) |
| Alpha-Beta Blockers | 71,725 (14.8%) |
| Vasodilators | 60,538 (12.5%) |
| Thiazide Diuretic | 43,925 (9.1%) |
| Potassium-Sparing Diuretics | 41,967 (8.6%) |
| Central-Acting Agents | 40,477 (8.3%) |
| Alpha Blocker | 8,420 (1.7%) |
| Aldosterone Antagonists | 435 (0.1%) |
| Renin Inhibitors | 58 (<0.1%) |

RRs were estimated using a modified Poisson regression model for having hypertension medication orders (Figure 1). RRs ranged from 0.91 to 1.09. The strongest positive association with medication orders was observed for hyperlipidemia medication use 1.09 [95% CI: 1.08, 1.09]), followed by diabetes medication use (RR = 1.05 [1.05, 1.06]) and (RR = Black or African American race (RR = 1.05 [1.04, 1.05]). Additional factors associated with a higher likelihood of treatment included Asian race (RR = 1.03 [1.03, 1.04]), increasing age (RR = 1.03 [1.03, 1.03]) and Hispanic or Latino ethnicity (RR = 1.02 [1.01,1.02]). In contrast, lower likelihoods of treatment were observed for other payer types (RR = 0.91 [0.91, 0.92]), diabetes diagnoses (RR = 0.95 [0.95, 0.95]), and hyperlipidemia diagnoses (RR = 0.98 [0.97, 0.98]).

**Figure 1.**
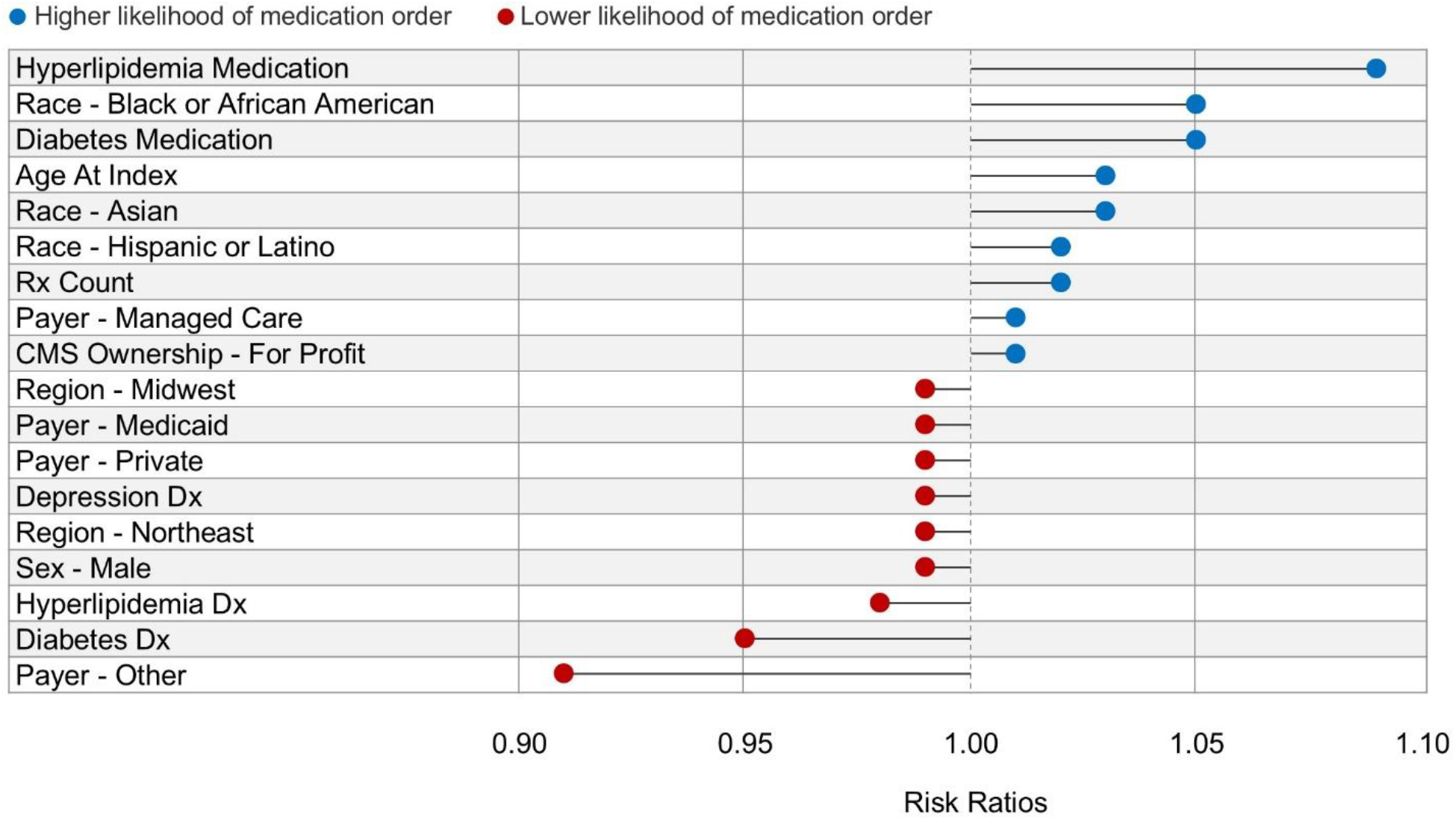
Summary of risk ratios. Abbreviations: CMS=Centers for Medicare and Medicaid; D=diagnosis; Rx=prescription. Note: The figure presents all variables in the model with risk ratios greater than or less than 1.00 with a p<0.001.

## Discussion

This study examined rates of pharmacological treatment and factors associated with antihypertensive treatment ordered among a large, national cohort of SNF residents in the United States. We found that 87.7% of residents diagnosed with hypertension had orders for pharmacologic therapy consistent within medication classes recommended by International Society of Hypertension, with calcium channel blockers and beta blockers as the most commonly prescribed medication classes. Residents prescribed diabetes or hyperlipidemia medications had a higher likelihood of having antihypertensive medication orders, while those with a diagnosis of diabetes or hyperlipidemia and those with other payer types had a lower likelihood.

The proportion of LTC residents receiving antihypertensive medication in this study is similar to recent national estimates, which indicate that treatment rates among older adults in the U.S. remain high, but not universal. ^13^ Despite advances in hypertension management and updated guidelines, gaps in care persist within SNF settings. Polypharmacy, clinical complexity, and competing priorities may contribute to undertreatment or therapeutic inertia. The predominance of beta blockers and calcium channel blockers in this study, despite guideline caution for first-line use of beta blockers in uncomplicated cases, highlights the health complexity of LTC residents.^10^ Disparities in hypertension treatment by race, region, and facility type were evident. Black or African American residents had higher odds of receiving pharmacological therapy compared to White residents. This may be due to the earlier onset and increased severity of hypertension in black individuals compared to white individuals.^14^ These findings underscore the need for targeted interventions to address both individual- and system-level determinants of care, as persistent inequities in hypertension management may increase cardiovascular risk and adverse outcomes in frail older adults. ^7^

Our results also revealed that residents prescribed hyperlipidemia or diabetes medications were more likely to receive antihypertensive treatment, whereas those with a diagnosis of diabetes or hyperlipidemia were less likely to be treated. This pattern likely reflects differences in care engagement, whereby residents actively managed with chronic medications may have more consistent clinical oversight, increasing the likelihood of concurrent hypertension treatment.^15^ In contrast, residents with untreated conditions may represent a subgroup with lower overall treatment intensity suggesting a broader gap in care. These residents may represent a blind spot in the SNF environment that may require individualized management and targeted monitoring. ^16^

Several limitations should be considered. The cross-sectional nature of the study limits the observed associations, and residual confounding, including unmeasured clinical factors and blood pressure values. Additionally, treatment patterns were assessed at a single point in time, precluding evaluation of residents’ prior treatment trajectories and other clinical circumstances that may shape prescribing decisions. The analysis was further limited to pharmacologic treatment and did not capture non–medication interventions such as dietary management or physical activity. Finally, the findings may not be fully generalizable to settings outside SNFs or to healthcare systems with different prescribing practices and care models. Despite these limitations, this study leverages a large, real-world EHR dataset and provides important insights into current patterns of hypertension care in LTC.

## Conclusions and Implications

In this large, national SNF cohort, while most SNF residents with hypertension were prescribed pharmacologic treatment, meaningful gaps and disparities remain. Medication orders were less common among residents with diabetes or hyperlipidemia diagnoses and those with certain payer types, while residents already receiving chronic disease medications were more likely to be treated. These findings suggest differences in care engagement and overall treatment intensity. The predominance of beta blockers and calcium channel blockers alongside these clinical gaps indicate that access to recommended therapies remains uneven across clinical and demographic groups. These results underscore the need for targeted strategies to strengthen hypertension management, support equitable prescribing, and enhance access to evidence–based therapies within SNF settings.

## Data Availability

Data produced in the present study are available upon reasonable request to the authors.

## Acknowledgements

We would like to thank Jody Long, MSN, MBA and Steve Buslovich, MD, CMD for their collaboration and their clinical input provided for this manuscript. We would also like to thank Michelle Sweeny, Aaron Norfolk, and Kaitlyn Riddell for providing their perspectives. This research was supported by PointClickCare Life Sciences and McMaster University. McMaster University Library provided journal article access.

